# An explainable AI approach for discovering social determinants of health and risk interactions for stroke in patients with atrial fibrillation

**DOI:** 10.1101/2023.02.27.23286470

**Authors:** Raquel M. Zimmerman, Edgar J. Hernandez, W. Scott Watkins, Nathan Blue, Martin Tristani-Firouzi, Mark Yandell, Benjamin A. Steinberg

## Abstract

Atrial fibrillation (AF) leads to significant morbidity and mortality, which is primarily related to stroke despite effective stroke prevention therapies. There remains a critical need for personalized, socially aware, equitable stroke risk prediction among patients with AF to enable optimal implementation of contemporary stroke-prevention therapies. In this brief report, we leverage innovative computational tools and high-quality, extensive data (1.8 m patients, augmented with social determinants of health information) to demonstrate the ability of a unique, explainable AI approach to improve the accuracy and equity of stroke risk prediction. Current risk stratification approaches are blind to social determinants of health and fail to adjust for unique contributions and interactions of variables upon stroke risk. In contrast, our results indicate that social determinants of health can be important modifiers of clinical variables and ultimately stroke risk. We hope that this analysis can provide evidence to drive better, more personalized, and equitable stroke risk stratification and prevention for patients with AF in the future.

## Main Text

Stroke remains the primary source of morbidity and mortality associated with atrial fibrillation (AF), despite major advances in prevention. While effective stroke-prevention strategies are available, optimal implementation of these treatments is limited by (1) rudimentary stroke risk stratification tools (i.e., CHADS_2_-VA_2_Sc), and (2) disparities in care and outcomes of AF. Over 150,000 yearly strokes in the US occur in patients with AF.^1^ Many of these occur among patients with AF who are misclassified as low-risk or fail to receive appropriate therapies due to healthcare disparities. Thus, there remains a critical need for personalized, socially-aware, and equitable stroke risk prediction for patients with AF. A recent NIH Workshop Report emphasized the need for computational approaches that include social determinants of health to eliminate the inequities in healthcare delivery and outcomes in AF.^2^ Here we demonstrate that explainable AI can fulfill this need.

We used data from the University of Utah Hospital Electronic Health Record repository comprising over 1.8 million patients to identify and analyze factors associated with AF and stroke. Standard terminologies and coding systems for healthcare, including ICD/SNOMED (diagnoses, clinical observations, and observation-qualifiers), CPT (procedures), RxNORM (medications), and LOINC (labs) were included as input variables. Leveraging modern, high-powered computational workflows, we selected features for analysis with a novel Poisson binomial-based approach to comorbidity discovery (PBC).^3, 4^ We then explored the association between clinical and social determinants of health variables and stroke in patients with AF by implementing a Probabilistic Graphical Model (PGM) AI technique. We use the multimorbidity network derived from this technique to understand the landscape of stroke risk and social determinants of health in patients with AF.

The overall patient population is shown in **Table 1**, consistent with a tertiary care AF cohort. In Figure 1, risk factors such as diabetes and hypertension do not yield similar risks of stroke (1.32x and 1.92x, respectively), though they are accorded the same value when determining CHADS_2_-VA_2_Sc score (1 point each). Moreover, among patients with a CHADS_2_-VA_2_Sc score in the treatment ‘Twilight’ zone (∼2), there is wide variability in relative risk (1.32x-2.26x). These results also demonstrate that SDoH, such as race, ethnicity, and insurance status can impact stroke risk among patients with AF (**Figure 1, Panel B**). For example, patients of American Indian or Pacific Islander ancestry with Medicaid have a greater than 50% increased risk of stroke compared with Hispanic patients with Medicaid. Finally, these results demonstrate that SDoH are important modifiers of clinical variables and ultimately stroke risk. Thus, there exist subgroups of patients for whom certain SDoH factors are critical for accurate risk stratification.

**Table 1:** Baseline characteristics of patients included in this study (N =16658).

| **Variable** | **Mean (SD) or n (%)** |
| --- | --- |
| Age | 71.18 (14.48) |
| Female | 7166 (0.43) |
| Ethnicity |  |
| Hispanic | 794 (0.05) |
| Non-Hispanic | 15376 (0.92) |
| Unknown | 488 (0.03) |
| Ancestry |  |
| Black | 178 (0.01) |
| White | 14837 (0.89) |
| Asian | 218 (0.01) |
| American Indian, Alaska Native | 127 (0.01) |
| Native Hawaiian, Pacific Islander | 206 (0.01) |
| Unknown | 1096 (.07) |
| Payer Type |  |
| Commercial | 4656 (0.28) |
| Medicaid/Self Pay | 971 (0.06) |
| Medicare | 10403 (0.62) |
| Unknown | 628 (0.04) |
| Comorbidities |  |
| Hypertension | 13483 (0.81) |
| Diabetes Mellitus | 6340 (0.38) |
| Vascular Disease | 6387 (0.38) |
| Congestive Heart Failure | 6423 (0.39) |
| Stroke | 2966 (0.18) |

**Figure 1.**
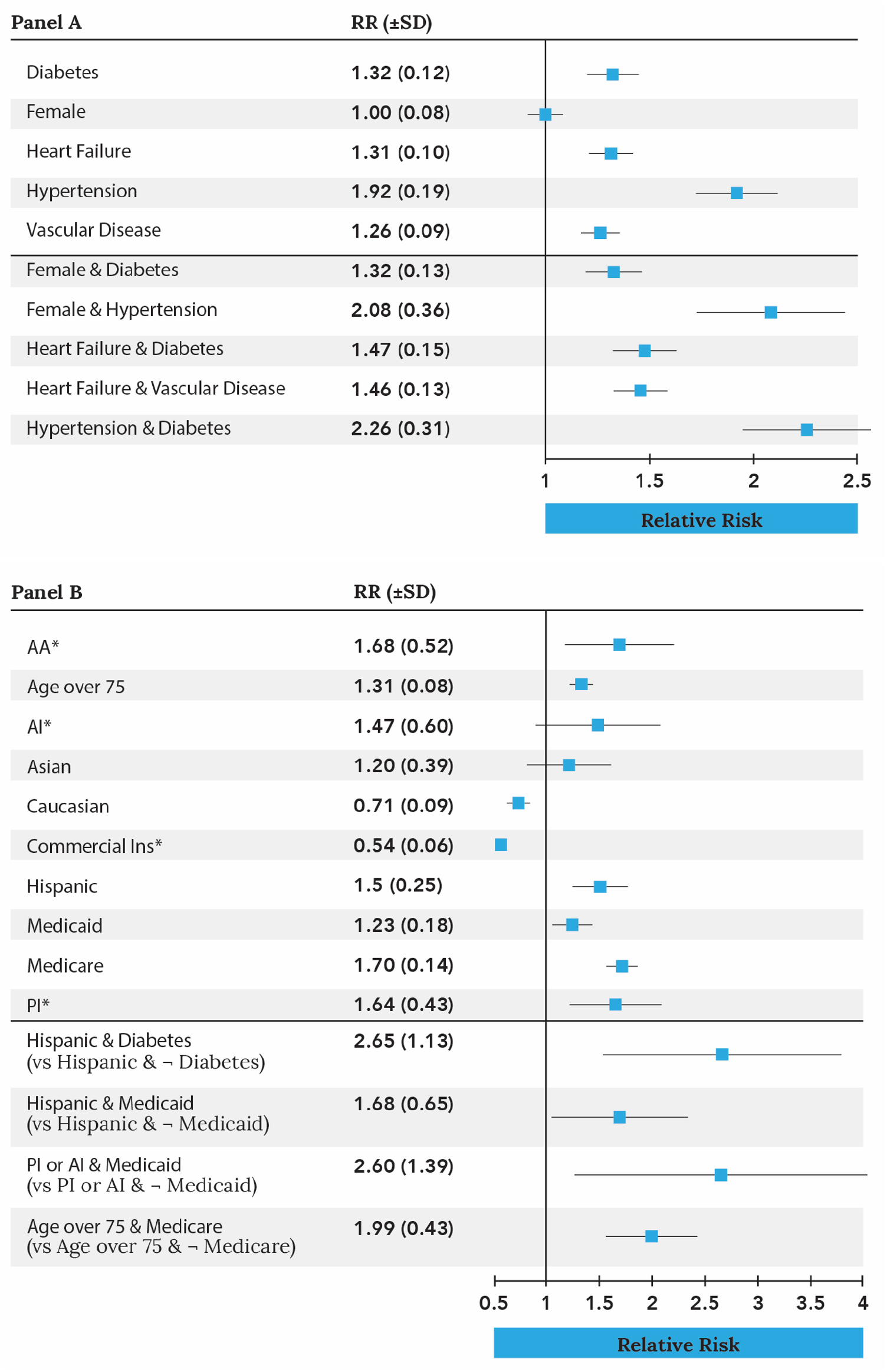
PGM-Based Relative Risks for Stroke among University of Utah Hospital Patients with Atrial Fibrillation. Panel A: Risk for various CHADS_2_-VA_2_Sc factors (top half all represent CHADS_2_-VA_2_Sc scores of 1; bottom scores of 2). Panel B: Impact of selected social determinants of health on stroke risk.

Our results highlight the limitations of current risk stratification approaches, which are blind to social determinants of health and fail to adjust for unique contributions and interactions of variables upon stroke risk. Our methodology moves beyond static scores, providing explainable, personalized stroke risk assessment while simultaneously addressing inequities of care. While the inclusion of self-described race and ethnicity poses challenges and has the potential to exacerbate inequities, a “color-blind” approach [a racial/ethnic-agnostic approach] may worsen prediction. In this context, it is likely to exacerbate inequities by under-estimation of risk in patients whose true risk would otherwise justify intervention.^5, 6^ The context-specific complexities of integrating these factors continue to require careful attention and further exploration in order to optimize equitable care delivery. Our analyses also demonstrate how explainable AI approaches can be used to provide personalized and equitable risk assessment strategies. We hope that future studies will further refine and improve this approach, discover and incorporate additional risk factors, and test implementation strategies for more personalized, precise stroke risk assessment among patients with AF.

## Data Availability

This data contains protected health information and cannot be shared without IRB approval.

